# Complications and treatment of hypercalciuria in patients affected by Familial Hyperkalemic Hypertension (FHHt)

**DOI:** 10.1101/2024.03.09.24303922

**Authors:** Viola D’Ambrosio, Elizabeth R Wan, Gerlineke Hawkins-van der Cingel, Keith Siew, Olivia McKnight, Pietro Manuel Ferraro, Stephen B Walsh

## Abstract

**Background and hypothesis:** Gordon syndrome (also pseudohypoaldosteronism type II (PHAII) or Familial Hyperkalemia with Hypertension (FHHt)) is a genetic condition characterised by hypertension, hyperkalaemia, hyperchloraemic metabolic acidosis and hypercalciuria caused by an activation of the thiazide-sensitive sodium-chloride cotransporter (NCC, encoded by *SLC12A3*) in the distal convoluted tubule of the kidney. Thiazides rescue the electrolyte and metabolic abnormalities however, it is not known whether they decrease urinary calcium excretion, nephrolithiasis and low bone mineral density.

**Methods:** We examined a cohort of 11 patients with genetically confirmed FHHt. Biochemical, radiological, and clinical data was obtained in patients before and after thiazide treatment. All patients gave informed consent.

**Results:** Among the FHHt cohort 5 of the 11 patients were female. 7 patients had heterozygous pathogenic variants in *KLHL3*, 3 patients had variants in *WNK4*, and one had a variant in *WNK1*. At baseline, only 1 patient was hypertensive, all patients were hyperkalemic, whereas only 40% of patients had low serum bicarbonate and increased urinary calcium excretion. 50% of patients also had low bone mineral density (either osteopenia or osteoporosis) and 1 patient had bilateral nephrolithiasis. 6 patients were treated with thiazide diuretic and therefore were suitable for comparison between pre and post treatment biochemistry and imaging data.

While both serum and urinary biochemistry was completely reverted after thiazide treatment, bone mineral density had a worsening trend. 1 patient presented with bilateral nephrolithiasis after thiazide treatment.

**Conclusion:** We demonstrate that thiazide treatment normalizes serum and urinary biochemistry. Thiazide treatment therefore has clinical utility even if hypertension or hyperkalaemia are not problematic. According to our study, thiazide treatment does not seem to revert loss of bone mineral density, however, whether thiazides have an impact in nephrolithiasis is less clear and our results may require larger samples.

## Introduction

Familial Hyperkalemia with Hypertension (FHHt) (also called pseudohypoaldosteronism type II (PHAII) or Gordon syndrome) is an ultrarare monogenic condition characterised by hypertension, hyperkalaemia, hyperchloraemic metabolic acidosis, hypercalciuria, suppressed plasma renin and low aldosterone^1^.

Biochemically and clinically, this condition mirrors Gitelman syndrome, a rare tubular disorder characterised by normal/low blood pressure, hypokalemia, metabolic alkalosis, hypomagnesemia, hypocalciuria and elevated renin and aldosterone^2^.

Gordon syndrome is a genetic disease, caused by heterozygous mutations in *CUL3, KLHL3, WNK1* and *WNK4* genes (autosomal dominant [AD] mutations) or biallelic mutations in *KLHL3* (autosomal recessive [AR] mutations)^3^. Hypercalciuria has been described in association to Gordon syndrome^4^, especially in patients with *WNK4* mutations^5^ in whom low bone mineral density has also been described. Patients with *KLHL3* mutations tend to be hypercalciuric as well, although no clear evidence on bone mass is available^1^. Patients with *WNK1* mutations appear to be normocalciuric^6^, whereas there is no data on patients with *CUL3* mutations. There is evidence that thiazides revert the electrolyte abnormalities in these patients^7^. However, there are no data on whether thiazide treatment reduces stone burden or halt the loss of bone mineral density, two consequences of hypercalciuria and FHHt.

## Methods

The main aim of our study is to describe changes in serum and urinary biochemistry, stone burden and bone mineral density in a cohort of patients affected by FHHt across different disease-causing mutations and after thiazide treatment. We retrospectively collected demographic, biochemical and radiological data from 2014 to 2022 from a cohort of 11 genetically confirmed FHHt patients followed at the London Tubular Centre, Royal Free Hospital in London, UK. Hypertension was defined as systolic blood pressure (SBP) ≥ 140 mm Hg or diastolic blood pressure (DBP) ≥ 90 mm Hg, according to the most recent National Institute for Health and Care Excellence (NICE) criteria. Mean arterial blood pressure (MAP) was calculated as DBP + 1/3(SBP – DBP). Estimated glomerular filtration rate (eGFR) was calculated with the CKD-EPI_cr_ formula ^8^. Hyperkalemia was defined as serum potassium > 5.0 mmol/L, hypercalciuria was defined as urinary calcium/creatinine (uCa/Cr) > 0.7 mmol/mmol and low serum bicarbonate (HCO_3_^-^) was defined as HCO_3_^-^ ≤22 mEq/L. Bone mineral density was evaluated by dual energy X-ray absorptiometry (DXA). Osteopenia was considered as a T-score between –1 and –2.5 and osteoporosisas a T score < –2.5 at any site, according to the WHO guidelines. Stone burden was assessed through computed tomography of kidneys, ureters and bladder (CT KUB). The study had ethics committee approval (REC 05/Q0508/6). Data were analyzed using non-parametric tests as appropriate (Prism GraphPad 8.0.2). This study was assessed by the Health Research Authority National Research Ethcis Service Ethics Committee of London-Bloomsbury(reference 05/Q0508/6). Ethical Approval was given.

## Results

We included 11 genetically confirmed FHHt patients (F:M=5:6), 7 with *KLHL3* mutation, 3 with *WNK4* mutation and 1 patient had *WNK1* mutation. Our cohort included 7 unrelated families. Baseline characteristics are shown in **Table 1**. All patients were young (median age 41) and with preserved kidney function (median eGFR 95 mL/min/1.73 m^2^). Only 1 patient (16%) was hypertensive pre-treatment. Of note, all patients were hyperkalemic at baseline (median sK^+^ 5.4 mmol/L), 40% of patients had low serum bicarbonate (median sHCO_3_ ^-^ 22.4 mEq/L) and 40% had increased urinary excretion of calcium (median uCa/Cr 0.6 mmol/mmol). Other parameters, including serum calcium, phosphate, alkaline phosphatase, magnesium and parathormone (PTH), were all in the normal range. At baseline, 50% of patients had low bone mineral density, 30% of patients with DEXA scans available were osteopenic, 1 patient was osteoporotic and 1 patient had bilateral nephrolithiasis (**Figure 1A**). Out of 11 patients, 6 were treated with a thiazide diuretic (bendroflumethiazide; mean dosage 2.75 mg/once daily) and were suitable for comparison of biochemical data, imaging and DEXA before and after thiazide treatment. Blood pressure did not significantly change pre and post treatment (**Figure 2**). With regards to biochemistry, thiazide diuretic normalised serum potassium and urinary calcium/creatinine ratio in all patients and in a statistically significant fashion (**Figure 3**). Serum bicarbonate improved as well although it did not reach statistical significance (**Figure 3**). All the other parameters did not significantly change after treatment and remained in the normal range. Out of the 6 patients treated with a thiazide diuretic, 5 had DEXA scans and 3 patients (60%) were osteopenic and 1 patient (20%) had DXA criteria for osteoporosis. Mean time between pre and post thiazide DXA scans was 12 months. Both osteopenia and osteoporosis were more prevalent in the lumbar spine. Pre and post thiazide comparison of total lumbar, L2 and femoral neck T-score showed a mild worsening trend, although it did not reach statistical significance (**Figure 4**). All patients treated with thiazide diuretic underwent a CT KUB; the patient that had bilateral nephrolithiasis pre thiazide (**Figure 1A**) showed no discernable kidney stones after treatment. However, another patient had bilateral nephrolithiasis even after treatment and normalisation of serum and urinary biochemistry (**Figure 1, panel B**).

**Table 1.**
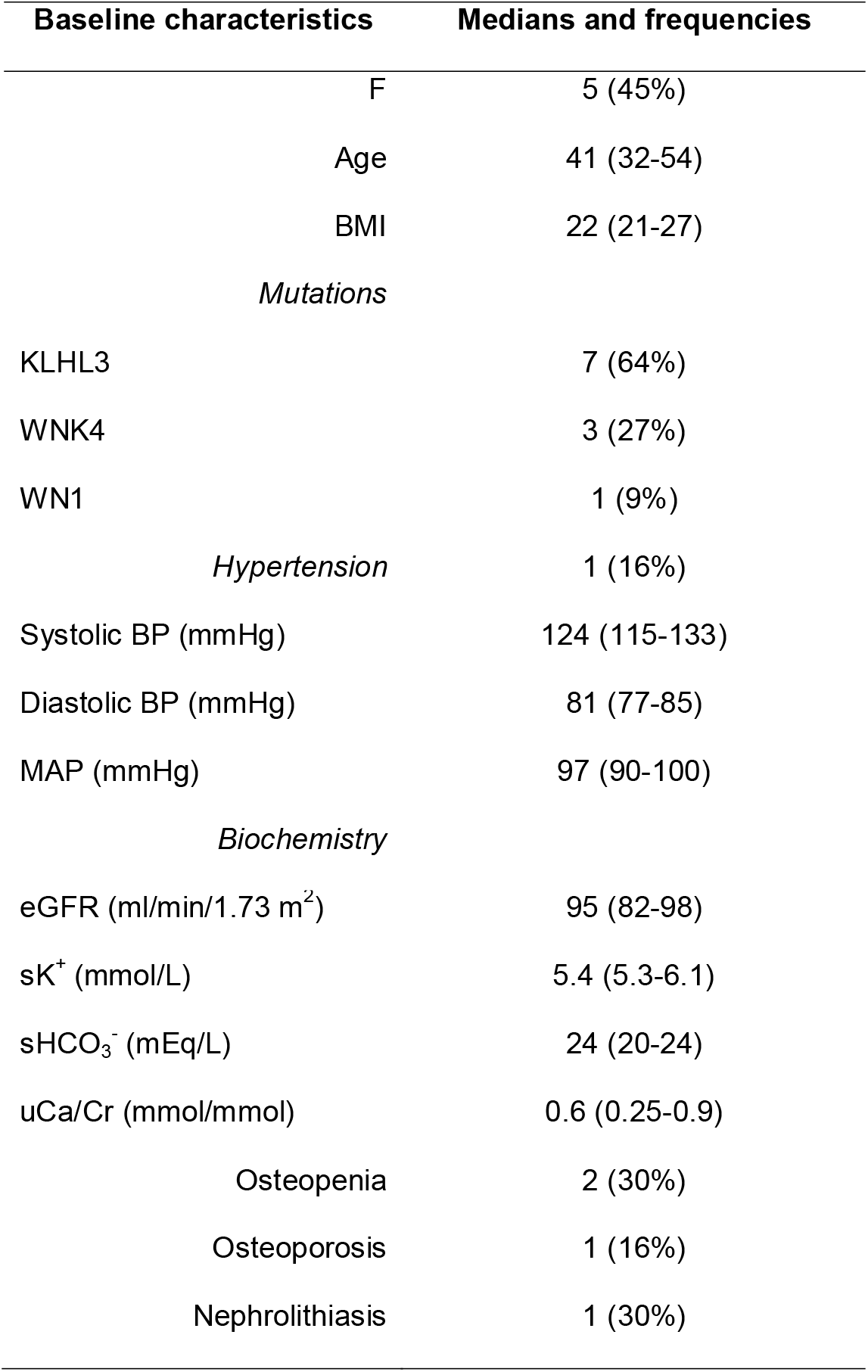
Baseline characteristics of the FHHt cohort. Continuous variables are reported medians and interquartile range (IQR) and categorical variables as frequencies and percentages. *BMI* body mass index; *KLHL3* kelch-like protein 3; *WNK4* and *WNK1* lysine deficient protein kinase 4 and 1; *BP* blood pressure; *MAP* mean arterial blood pressure; *eGFR* estimated glomerular filtration rate; *sK*^*+*^ serum potassium; *sHCO*_*3*_^*-*^ serum bicarbonate; *uCa/Cr* urinary calcium creatinine ratio.

**Figure 1.**
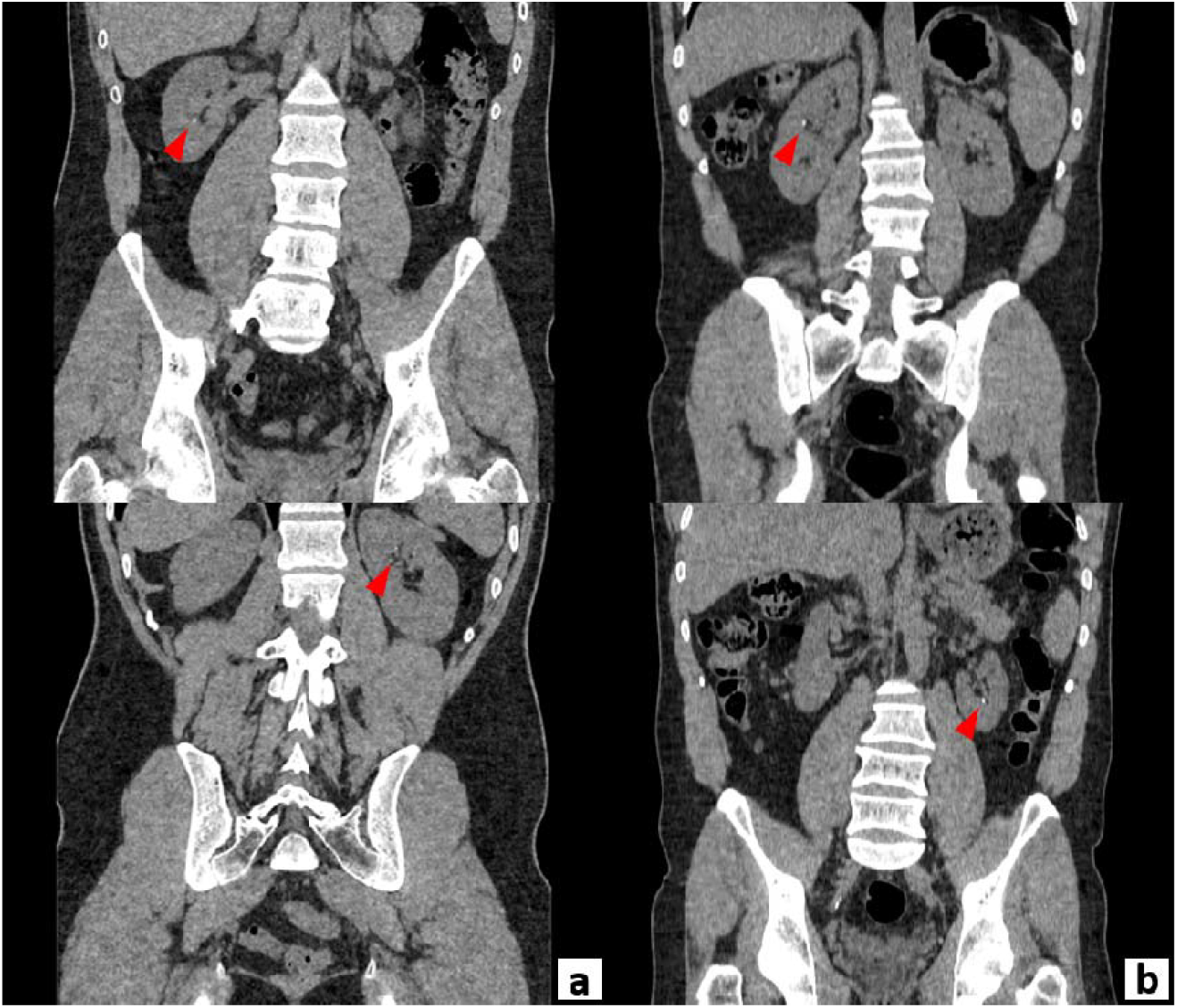
A Pre thiazide CT KUB showing bilateral nephrolithiasis consisting of a 2 mm calculus in the right lower pole and 1 mm calculus in the left upper pole (red arrowheads). B Post thiazide CT KUB in a different patient showing bilateral nephrolithiasis consisting of a 3 mm calculus in the interpolar region of the right kidney and a 4 mm calculus in the left lower pole (red arrowheads).

**Figure 2.**
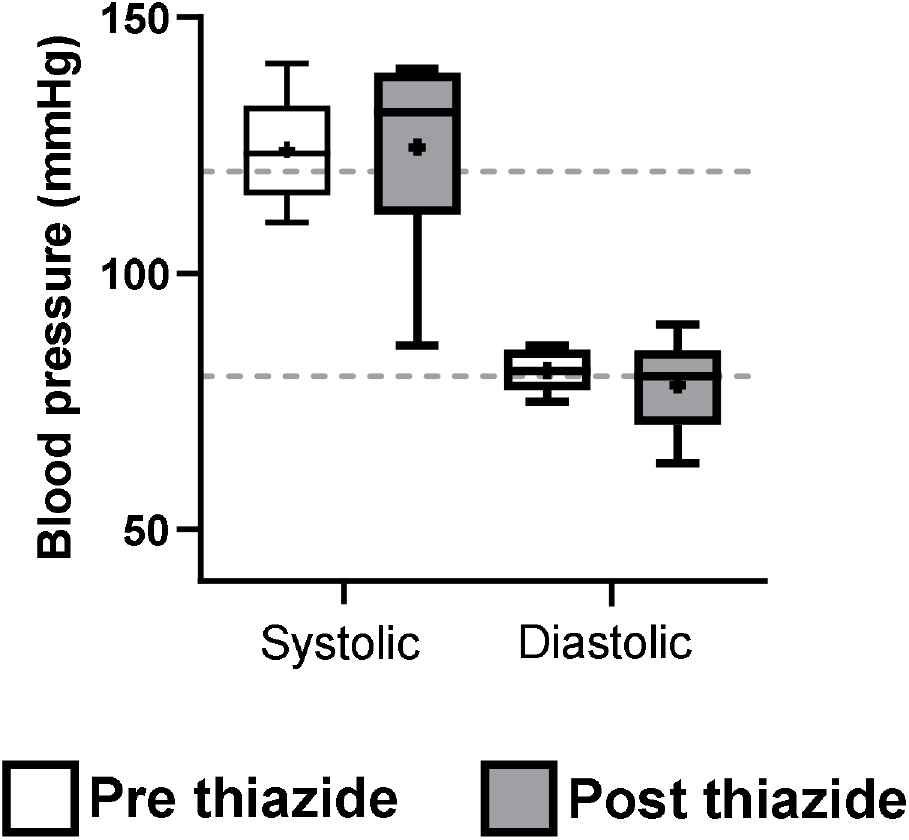
Comparison of blood pressure pre and post thiazide treatment. Box plots show median, mean (+), minimum and maximum (whiskers) and lower Q1 and upper Q3 quartile range (lower and upper limit of the box, respectively). Grey dotted lines represent normal systolic (120 mmHg) and diastolic (80 mmHg) blood pressure values.

**Figure 3.**
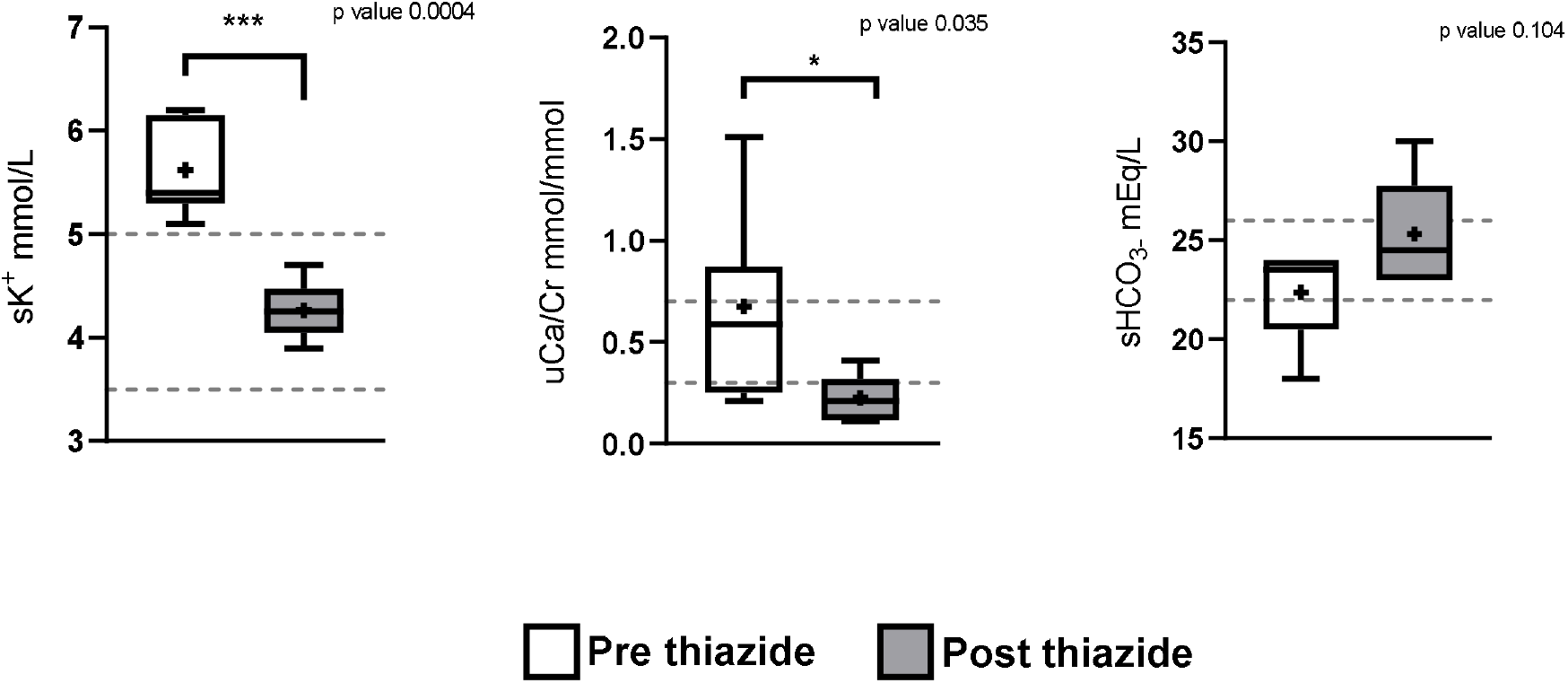
Comparison of biochemistry pre and post thiazide treatment. Box plots show median, mean (+), minimum and maximum (whiskers) and lower Q1 and upper Q3 quartile range (lower and upper limit of the box, respectively). Grey dotted lines represent upper and lower limit of the normal range. *sK*^*+*^ serum potassium; *sHCO*_*3*_^*-*^ serum bicarbonate; *uCa/Cr* urinary calcium creatinine ratio.

**Figure 4.**
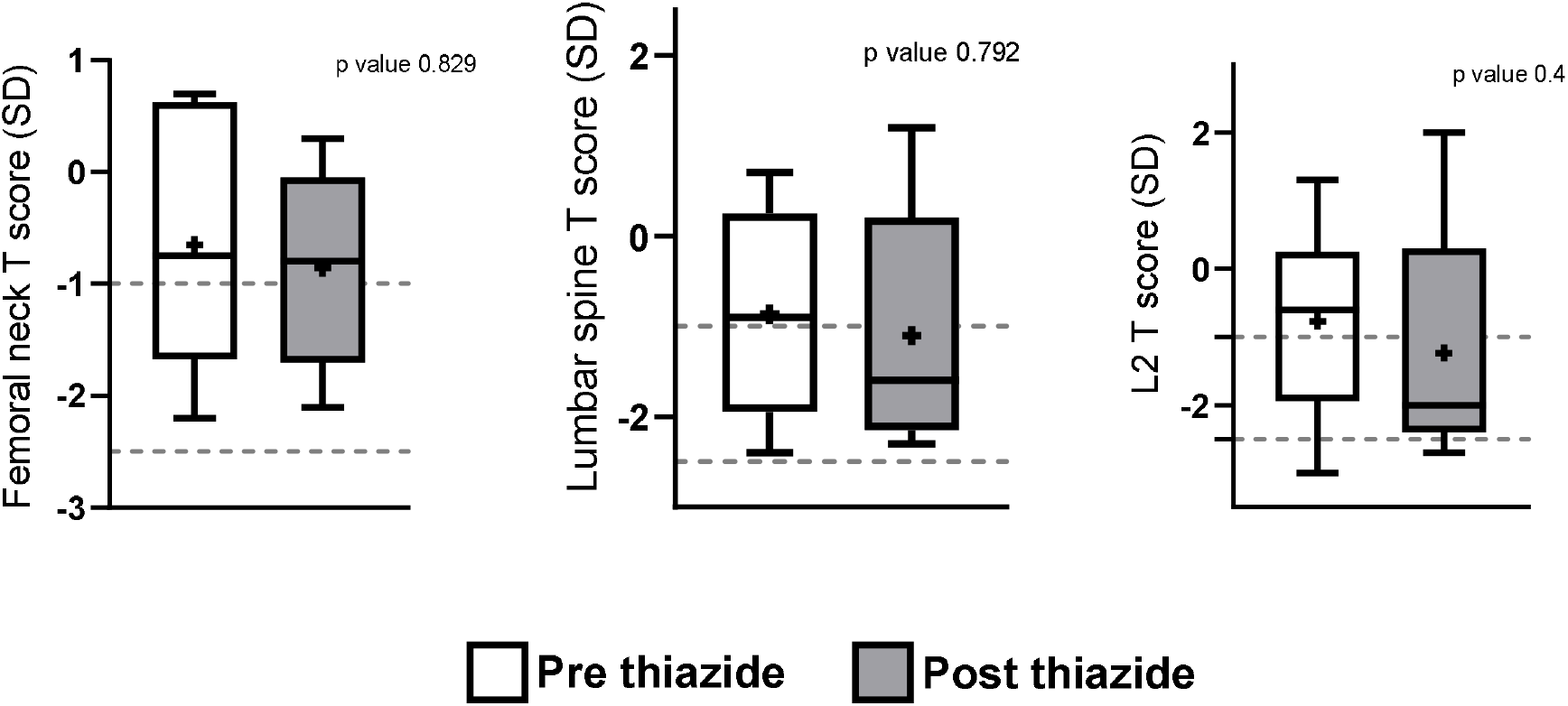
Comparison of lumbar spine, L2 and femoral neck T score pre and post thiazide treatment. Osteopenia was defined as a T score between -1 and -2.5; osteoporosis was defined as a T score < -2.5. Box plots show median, mean (+), minimum and maximum (whiskers) and lower Q1 and upper Q3 quartile range (lower and upper limit of the box, respectively). Grey dotted lines represent upper and lower limit of the osteopenia range.

## Discussion

FHHt was first reported by Paver and Pauline in 1964^9^ and then described by Richard Gordon and coworkers (1970) in patients that presented with hypertension and hyperkalemia, but normal glomerular filtration rate ^10–12^. Before knowing the underlying physiological mechanism, patients were given loop diuretics in order to control hypertension and hyperkalaemia, however, these drugs worsened the excretion of urinary calcium and consequently caused loss of bone mineral density and increased kidney stone burden. FHHt is characterised by over-activation of the thiazide-sensitive Na-Cl cotransporter (NCC) in the distal convoluted tubule (DCT). NCC activation is caused by dysregulation of the upstream serine/threonine kinase (importantly including the ‘with no lysine’ or ‘WNK’ kinases, WNK1 and WNK4) cascade that regulates NCC expression and phosphorylation. This leads to increased reabsorption of sodium and water, volume expansion, hypertension and therefore the compensatory normal-low circulating aldosterone levels and low renin levels. WNK4 also regulates the transient receptor potential V5 channel (TRPV5), a Ca^2+^ channel in the connecting tubule (CNT) that regulates calcium transport ^13^. TRPV5 is an apical transporter that mediates passive transcellular calcium transport in the DCT. WNK4 mutations downregulate TRPV5 causing decreased reabsorption of calcium in the DCT and therefore hypercalciuria. An additional potential mechanism could be that the intracellular increase in Na^+^ concentration causes increased intracellular concentration of Ca^2+^ by the increased activity of the basolateral Na/Ca exchanger (NCX1) ^14^. This could eventually lead to decreased activity of TRPV5 with consequent reduced Ca^2+^ reabsorption from the tubular lumen and hypercalciuria. Another possible mechanism that have been hypothesized is the decreased Ca^2+^ reabsorption in the upstream nephron^15^, more specifically in the proximal tubule as a consequence of compensatory reduced Na^+^ reabsorption^16^. Hypercalciuria is not a consistent finding in all the FHHt-causative mutations. In a case series published by Mayan *et al* ^1^ two families with *KLHL3* mutations were reported to have hypercalciuria in addition to all the typical clinical features of FHHt. In one family, the two index cases were two identical twins that were referred for short stature. All affected cases presented with hypercalciuria, although milder than the those with WNK4 mutations. Patients with *WNK1* mutations tend to be normocalciuric as described by Achard *et al* in a large family with *WNK1*-FHHt^6^. To date, there is no published data detailing whether patients with *CUL3* mutations are hyper or normocalciuric. Nephrolithiasis has been previously described in association with FHHt although it is not a common finding. The first case report was described in 1998, a 40-year-old man with monolateral nephrolithiasis incidentally diagnosed during a routine abdomen ultrasound (US) whose composition analysis after ESWL revealed the presence of both calcium oxalate and calcium phosphate^17^. In the family described by Mayan *et al*^18^, only one patient out of eight affected presented with nephrolithiasis prior to thiazide treatment. More recently, another case report described an adult male patient affected by FHHt with a history of nephrolithiasis during adolescence that required surgical removal^19^. Increased urinary excretion of calcium is a well-known risk factor for nephrolithiasis, therefore it is not surprising that these patients get stones^20^. Reduced bone mineral density is another feature of Gordon syndrome, although its prevalence and its definite etiology have not been fully elucidated. Mayan *et al* described a family with eight members affected by Gordon’s syndrome, whose DXA scans showed evidence of low bone mineral density^18^. In a Portuguese population study, a specific WNK4 mutation (R1204C) was found to be associated with decreased bone mineral density or ‘osteoporosis’ ^21^.

More specifically, the study was conducted on hypertensive and osteoporotic patients to assess the prevalence of WNK4 genetic variants and to hypotheses its possible role in both diseases. The R1204C mutation of WNK4, although very rare among the general population, was more prevalent in osteoporotic patients. More than one mechanism could contribute to loss of bone mineral density in these patients: 1. NCC is expressed by osteoblasts^22^. Inactivation of NCC, either in mouse models of Gitelman syndrome or via pharmacological inhibition (thiazide treatment), increases osteoblasts differentiation and activity leading to enhanced deposition of bone^23^. In Gordon syndrome, an over-activation of NCC could inhibit osteoblasts and favor bone reabsorption. 2. Hypercalciuria alone has been associated to reduced BMD^24^, 3. Elevated parathyroid hormone (PTH) or subclinical hyperparathyroidism secondary to hypercalciuria could have a role in increased bone reabsorption and release of calcium salts in the bloodstream to maintain serum calcium concentrations within normal range (in our cohorts, all patients had an available PTH record and they were all in the normal range). In one case report, hyperparathyroidism secondary to hypercalciuria was successfully treated with thiazides^25^. 4. Hypertension itself has been reported to be associated with reduced BMD^26^, 5. Metabolic acidosis is a well-known risk factor for osteomalacia/low BMD. Bone represents an important source of calcium salts that buffer acidemia at the expenses of bone remodeling. Indeed, short stature related to FHHt has been previously described^10,27^; this finding was confirmed in a study conducted by Farfel *et al* ^28^ in a family of 57 members, 30 of whom affected by FHHt with WNK4 mutations. Short stature is not a consistent finding in patients affected by FHHt nor in families with the same mutations.

Speculations about the possible underlying mechanisms have been made, but not yet proven. In our cohort of patients, none have short stature. As extensively reported, thiazides are the main treatment for FHHt as they rescue all of the electrolyte abnormalities. However, whether thiazides affect other clinical manifestations such as nephrolithiasis or low bone mineral density is not known. From our results, nephrolithiasis and low bone mineral density persist after thiazide treatment. To our knowledge this is the first study to systematically evaluate nephrolithiasis and mineral bone density in a well characterised FHHt cohort. Most patients included in the study were not related and, although our cohort was rather large for this ultra-rare disease, our study lacked genetic variability since most of the patients have either WNK4 or KLHL3 mutations.

## Conclusions

Nephrolithiasis and low bone mineral density are complications of Gordon’s syndrome. We propose that patients with a suspected or confirmed diagnosis of Gordon’s syndrome should undergo imaging to exclude the presence of kidney stones (ideally CT KUB) and a DXA scan to assess bone mineral density. A thiazide diuretic is the current optimal treatment for Gordon’s syndrome as it reverses the electrolyte and metabolic abnormalities, however whether thiazides do not reduce the risk of further kidney stones or revert low bone mineral density within the time frame studied here is not yet clear.

## Data Availability

All data produced in the present study are available upon reasonable request to the authors

## Data Availability Statement

The data that support the findings of this study are available from the corresponding author, SBW, upon reasonable request

## Acknowledgments

We thank the patients and their families for the constant support in research.

